# Implementing a Group Education Model for People with Type 2 Diabetes in a Socioeconomically Deprived Primary Care Population: A Quality Improvement Report

**DOI:** 10.64898/2026.11.04.26366897

**Authors:** Emily Bond, Jane Sennett, Kristin Veighey

## Abstract

People with type 2 diabetes mellitus (T2DM) receiving injectable therapies require ongoing education and support to optimise self-management and glycaemic control. Group education may offer a cost-effective approach that combines professional guidance with peer support. Homewell Practice serves a socioeconomically deprived population in Hampshire with significant health inequalities. Many patients receiving insulin or GLP-1 receptor agonist therapy had poor glycaemic control, frailty, or increased risk of hospital admission.

Traditional one-to-one consultations provided limited opportunities for peer learning and support. A 12-month quality improvement project was conducted between May 2022 and April 2023. Adults with T2DM receiving injectable therapy were invited to attend monthly community-based education sessions. Attendance data, HbA1c outcomes and patient feedback were collected and analysed descriptively. Patient-centred group sessions were delivered using adult-learning principles and peer discussion. Topics included insulin titration, hypoglycaemia management, nutrition, glycaemic targets and diabetes complications.

Fifteen patients attended at least one session. Improvements in glycaemic control were associated with repeated attendance. Mean HbA1c reduction was 12.4% in patients attending four or more sessions, compared with 5.6% for those attending three sessions and 2.4% for those attending two sessions. Patients attending only one session demonstrated a 1.8% increase in HbA1c. All questionnaire respondents (9/9, 100%) reported improved understanding of diabetes and found the sessions useful; 8/9 would recommend the programme. Peer support was identified as the most valued aspect of participation. Attendance remained lower than anticipated, particularly among patients with multiple comorbidities and mental health needs.

Group education for adults receiving injectable diabetes therapy was associated with high patient satisfaction and improved glycaemic outcomes among regular attendees. A hybrid model combining group education with individualised support may improve accessibility and engagement for higher-need patients.

## PROBLEM

Homewell Practice serves a socioeconomically deprived population within Havant, Hampshire, including residents of the Leigh Park estate. The practice population of around 15,000 patients supports approximately 1,106 patients with type 2 diabetes, including 291 receiving injectable therapies such as insulin and GLP-1 receptor agonists. Many of these patients have poor glycaemic control, frailty, complex comorbidity and an increased risk of hospital admission.

The Indices of Multiple Deprivation (IMD) show that Leigh Park has some of the most deprived wards in England^1^. Battins ward has the highest percentage of residents in the Borough who have ‘‘no qualifications’’ and who regard themselves in ‘‘very bad health’’. Additionally, Warren Park ward has the highest percentage of residents in the Borough who are long-term unemployed or who have never worked^1^.

Diabetes UK released a report in 2009 titled ‘Diabetes in the UK 2009: Key statistics on diabetes’^2^. The report showed that deprivation is strongly associated with higher levels of obesity, physical inactivity, unhealthy diet, smoking and poor blood pressure control, all of which are linked to the risk of developing Type 2 diabetes and the risk of serious complications amongst those already diagnosed with both Type 1 and Type 2 diabetes. We also know that people living in more socio-economically disadvantaged neighbourhoods have higher rates of almost all of the known underlying clinical risk factors that increase the severity and mortality of COVID-19.^3^

At the time of the initial service proposal in 2021, the impact of the global pandemic had significantly adversely affected the delivery of diabetes care for primary care teams throughout the UK. Many patients were now overdue annual screening reviews, which the authors felt put them at great risk of higher mortality and hospital admission with COVID-19, and the development of, or progression of, long term diabetic complications.

Following disruption to routine diabetes care during the COVID-19 pandemic, many patients were overdue annual review and there were concerns regarding worsening glycaemic control and progression of diabetic complications. It was felt that traditional one-to-one consultations provided limited opportunities for peer learning and support.

The aim of this quality improvement project was to develop and evaluate a community-based group education programme for adults with type 2 diabetes receiving injectable therapies.

Specific objectives were to improve patient knowledge, confidence in self-management, understanding of glycaemic targets and hypoglycaemia management, and ultimately improve glycaemic outcomes.

## BACKGROUND

Structured diabetes self-management education is recommended by NICE as a core component of management for people with type 2 diabetes^4^. Previous studies have demonstrated improvements in glycaemic control, self-management behaviours and patient confidence following diabetes self-management education programmes^5^. Group-based education may additionally provide opportunities for peer support, shared learning and social reinforcement of behaviour change^4^. However, engagement with structured education is often lower amongst socially disadvantaged populations and people living with multiple long-term conditions^3^.

Group-based educational interventions offer a potential means of improving access to diabetes education whilst facilitating peer support and shared learning. In addition to receiving information from healthcare professionals, participants can learn from the experiences of others living with diabetes, helping to normalise challenges, exchange practical coping strategies and reinforce behavioural change^4,7^. Previous studies have demonstrated that group education can improve clinical outcomes and patient confidence whilst providing a cost-effective approach to supporting larger numbers of patients.

Theoretical models of adult learning suggest that adults learn most effectively when educational content is relevant to their own experiences, builds upon existing knowledge and encourages active participation^8,9^. Person-centred group discussions therefore have the potential to enhance engagement and support individuals in applying new knowledge to their daily diabetes management. Such approaches may be particularly valuable for people receiving injectable therapies, who may face additional challenges relating to medication management, blood glucose monitoring and concerns regarding hypoglycaemia.

Despite growing evidence supporting structured education, uncertainty remains regarding the most effective methods of delivering programmes within routine primary care, particularly in communities affected by social deprivation where attendance and engagement may be challenging. Consequently, there is a need to evaluate innovative, community-based approaches that combine professional education with opportunities for peer support and shared learning.

## MEASUREMENT

The primary outcome measure for this project was change in glycaemic control as assessed by HbA1c. HbA1c was selected as it represents an established indicator of diabetes control and is routinely measured in primary care. Given the educational focus of the intervention, secondary outcome measures included participant attendance, self-reported confidence in diabetes self-management, understanding of diabetes, and overall satisfaction with the programme.

Baseline assessment involved identifying all adults with type 2 diabetes receiving injectable therapies within the practice population. Patients were initially prioritised according to predefined risk criteria including suboptimal glycaemic control (HbA1c ≥75 mmol/mol), frailty and mixed insulin use, and increased risk of hospital admission. Demographic and clinical characteristics were obtained from routine electronic health records. Clinical outcome data were collected through routine diabetes reviews undertaken as part of standard care. HbA1c values before and following participation in the programme were extracted from electronic patient records and analysed descriptively. As this was a service improvement initiative delivered within routine clinical practice, the timing of follow-up blood tests was not standardised across all participants. Patient-reported outcomes were collected using anonymous post-session questionnaires immediately following attendance at educational sessions.

Participants were asked to rate their understanding of diabetes, confidence in self-management, satisfaction with the educational content, and whether they would recommend the programme to others. Free-text comments were also invited to capture individual experiences and identify opportunities for service improvement. Attendance records were maintained for each session to assess engagement with the programme and explore whether frequency of attendance was associated with differences in clinical outcomes. Data were analysed descriptively owing to the small sample size and service evaluation design.

The intention was to evaluate the feasibility, acceptability and potential impact of a community-based group education model within routine primary care rather than to demonstrate definitive effectiveness.

## DESIGN

A community-based group education programme was developed for adults with type 2 diabetes receiving injectable therapies within a large primary care practice serving a socioeconomically deprived population. The intervention was designed in response to concerns regarding poor glycaemic control, increasing clinical complexity and the limited opportunity for peer support within traditional one-to-one consultations. The programme was designed and delivered by a multidisciplinary team led by a Diabetes Specialist Nurse (DSN) Jane Sennett and a Clinical Pharmacist with a specialist interest in diabetes (Emily Bond), and supported by nursing staff, student nurses and administrative colleagues. Prior to this programme, an informal group clinic had been facilitated around April to December 2019 before the COVID-19 pandemic. These clinics were led by our diabetes lead Jane Sennett and did not record any outcomes or invitation structure, but informal and anecdotal feedback from these sessions from patient attendees had been used as the basis of patient participation and involvement (PPI) considerations. Patient involvement is increasingly recognised as an important component of service improvement, with evidence suggesting that involving people living with diabetes in the design and development of services can improve care and patient experience^10^.

Educational content was informed by established principles of diabetes self-management education and adult learning theory, with an emphasis on active participation, shared learning and person-centred discussion. Rather than delivering a fixed educational curriculum, sessions were structured to encourage participants to identify topics that were most relevant to their own experiences and learning needs. Sessions were delivered in a local community venue selected for its accessibility, familiarity and non-clinical environment. The intention was to create a welcoming setting that reduced some of the barriers associated with traditional healthcare environments and encouraged open discussion between participants. Educational sessions incorporated both professional input and facilitated peer discussion, allowing participants to share experiences, practical strategies and challenges associated with living with diabetes and managing injectable therapies. Core topics included glycaemic targets, blood glucose monitoring, hypoglycaemia recognition and management, lifestyle modification, medication adherence, diabetes complications and cardiovascular risk reduction. However, session content remained flexible in order to respond to participant questions and priorities. This approach was intended to increase engagement and support the application of learning to individual circumstances.

The programme was funded through educational grants from industry partners, enabling venue hire, educational resources and programme delivery. To maintain independence, content development and delivery remained under the control of the clinical team. The underlying theory of the intervention was that combining structured diabetes education with opportunities for peer support would improve participants’ knowledge, confidence and self-management capability. It was anticipated that increased engagement with diabetes self-management behaviours would contribute to improvements in glycaemic control whilst providing a scalable and sustainable model of care that could be delivered within routine primary care services.

## STRATEGY

The planning stage commenced in February 2022, with the vision of monthly face-to-face educational sessions to be delivered in a community venue from May 2022 to April 2023. Initial invitations were targeted towards individuals considered to be at highest risk of adverse diabetes outcomes:

- Patients identified as moderately or severely frail who were currently using mixed insulin products. This criterion was selected because of the increased risk of adverse events in this group compared with individuals living with moderate or severe frailty who were receiving intermediate or basal insulin only.^11^
- Patients using injectable diabetes therapy (insulin or GLP-1 receptor agonists) with a most recent HbA1c result of **75 mmol/mol or higher**.
- Patients with a **QAdmissions score of 20% or greater**^12^.

This approach was adopted on the assumption that individuals with the greatest clinical need would derive the greatest benefit from enhanced education and support. Initially patients fitting all three criteria were invited, then patients fitting 2 out of the 3 criteria were invited.

However, attendance during the early stages of the programme was lower than anticipated.

**Table 1.** Initial prevalence data and criteria stratification.

|  | Number of patients at Homewell Practice |
|---|---|
| Patients with type 2 diabetes | 1106 |
| Patients with T2DM prescribed insulin therapy | 187 |
| Patients with T2DM prescribed GLP-1 therapy | 136 |
| Patients with T2DM prescribed either insulin or GLP-1 therapy or both concomitantly | 291 |
| Moderate/severe frailty using mixed insulin | 38 |
| Any injectable therapy HBA1C ≥ 75mmol/mol | 97 |
| QAdmissions ≥ 20% | 65 |
| Patients fitting all three criteria | 7 |
| Patients fitting any two of the criteria | 50 |
| Patients fitting any one criterion | 125 |

Following review of attendance patterns, eligibility criteria were widened in June 2022 to include patients meeting a single risk criterion. This change aimed to increase accessibility whilst maintaining a focus on patients most likely to benefit from additional support.

Although attendance improved modestly, engagement remained below expectations. It became apparent that restricting invitations to higher-risk individuals may have limited participation and reduced opportunities for peer interaction within the groups.

In August 2022, recruitment criteria were further expanded to include all adults with type 2 diabetes receiving injectable therapies within the practice. This broader approach was intended to maximise accessibility and create a more diverse learning environment in which participants could share a wider range of experiences and self-management strategies. This recruitment strategy generated the greatest level of engagement and was subsequently maintained for the remainder of the programme. Throughout the project, educational sessions remained flexible and responsive to participant needs. While key topics were identified in advance, facilitators encouraged discussion of issues raised by attendees and adapted session content accordingly. Participant feedback consistently highlighted the value of peer support and shared learning, reinforcing the decision to maintain a discussion-based format rather than delivering a rigid educational curriculum. Practical aspects of programme delivery were also reviewed continuously, including venue suitability, methods of patient invitation and session organisation. These adaptations were intended to improve participant experience and support the long-term sustainability of the programme within routine primary care.

The principal learning from the implementation process was that broader eligibility criteria and a flexible participant-led approach appeared to promote greater engagement than a highly targeted recruitment strategy. The iterative modifications undertaken throughout the project helped shape a model that was acceptable to participants while remaining deliverable within existing primary care resources.

## RESULTS

### Participant Characteristics

**Table 2.** A summary of the demographics of the attendees and number of sessions attended.

| **Measure** | **Value** |
|---|---|
| Unique participants | 15 |
| Female participants | 8 (53.3%) |
| Male participants | 7 (46.7%) |
| Number of sessions delivered | 12 |
| Average attendance per session | 6.4 |
| Attendance range | 4-12 participants |
| Attended 1 session | 5 (33.3%) |
| Attended 2 sessions | 1 (6.7%) |
| Attended 3 sessions | 3 (20%) |
| Attended ≥4 sessions | 6 (40%) |
| Attended all 12 sessions | 0 |

Questionnaire data were available for 9 of the 15 attendees. HbA1c follow-up data reflected routine clinical testing and were therefore not available at standardised intervals for all participants.

### Summary of cohort changes during the service

Recruitment criteria evolved throughout the project. Initially, invitations were limited to patients meeting two or more risk criteria. In June 2022 the programme expanded to include patients meeting a single criterion, and from August 2022 all adults receiving injectable therapy for T2DM were eligible. This was because the expected uptake of invitation was far less than anticipated.

### Attendance Patterns

The highest attendance occurred in the final session (April 2023) with 12 participants, while the lowest was in August 2022 with only 4 attendees. Most sessions averaged 4-6 participants, with supporters (carers or spouses) occasionally attending.

### HBA1C Outcomes

**Table 3.** Summary of mean HBA1C changes against frequency of attendance.

| Attendance | Mean HBA1C change |
|---|---|
| One session | +1.8% |
| Two sessions | -2.4% |
| Three sessions | -5.6% |
| Four or more sessions | -12.4% |

### Patient reported outcomes

Nine out of 15 attendees completed anonymous paper evaluation questionnaires, revealing overwhelmingly positive feedback. All respondents found the sessions useful, with 4 rating them as “very useful” and 5 as “somewhat useful.” No participants reported the sessions as uncertain or not useful. The most valued educational theme was sharing experiences with other people living with type 2 diabetes, identified by 8 out of 9 respondents. Other highly appreciated topics included understanding HBA1C and blood sugar levels (6 responses), treating hypoglycaemia (6 responses), injection techniques (5 responses), and the effect of different foods on blood glucose levels (5 responses). Less popular topics included lifestyle advice, blood pressure management, and cholesterol education. When asked about factors that might improve attendance, most patients were satisfied with current arrangements, with only one requesting timing changes and another suggesting more advance notice for work shift adjustments.

- All 9 survey respondents reported improved understanding of their condition, while 7 noted better understanding of how food and drink impact blood sugar control. Six patients reported actual blood sugar improvements, and smaller numbers reported benefits including reduced hypoglycaemia frequency and improved understanding of diabetic complications.
- Eight out of 9 patients would recommend group sessions to others with diabetes or long-term conditions. Regarding overall impact, 6 patients reported extremely positive effects on their diabetes management, while 3 reported somewhat positive effects. No participants reported negative impacts or zero change.

The qualitative feedback revealed high satisfaction with the educational content and delivery. Patients appreciated the knowledgeable staff, regular medication adjustments, and particularly valued hearing about others’ experiences and problem-solving approaches.

Comments highlighted the importance of peer connection: “Good to see and hear other people’s experiences and problems” and “Important to know that others share your problem.” There was an emphasis on the value of peer support, knowledgeable staff, and practical diabetes management advice, with patients particularly appreciating the opportunity to learn from others’ experiences and receive regular medication adjustments in a supportive group environment.

## LESSONS AND LIMITATIONS

This service evaluation explored the feasibility and outcomes of a community-based group education programme for adults with type 2 diabetes receiving injectable therapies. Despite lower-than-anticipated attendance, participants who attended regularly demonstrated greater improvements in HbA1c than those attending fewer sessions. Patients consistently identified peer discussion and shared learning as the most valuable aspects of the intervention.

One of the strengths of this intervention was that it was developed and delivered within routine NHS primary care serving a population experiencing significant socioeconomic deprivation and health inequality. Many diabetes education studies are undertaken in research settings or specialist services, whereas this intervention reflects the realities of delivering care within a high-need community. The project specifically targeted individuals receiving injectable therapies, many of whom had poor glycaemic control, frailty or an increased risk of hospital admission.

Patient feedback consistently identified peer discussion as the most valuable component of the intervention. The group model appeared to allow participants to learn not only from healthcare professionals but also from others living with similar challenges, helping to normalise difficulties and facilitate practical problem-solving . Although the sample size was small, a clear relationship was observed between session attendance and improvements in HbA1c. Participants attending four or more sessions experienced substantially greater reductions in HbA1c than those attending fewer sessions, suggesting sustained engagement may be important in achieving clinical benefit^5^.

Despite positive outcomes for attendees, overall attendance was lower than anticipated, with only 15 patients participating over the 12-month period. The project attendance outcomes suggested that patients with the highest need for support—those with mental health conditions and multiple comorbidities—were less likely to engage in group settings, preferring individual consultations. Potential explanations include concerns regarding privacy, confidence in group participation, competing social priorities and practical barriers to attendance. These factors were not formally explored within this evaluation. There is also a suggestion that patients struggling with multiple social and health conditions may disengage from regular patient care in a standard setting.^13,14^

This resulting small sample size will have an impact on the perceived significance of the study; in addition to this limitation, there was also no control group. Invitation bias was mitigated by ensuring that all eligible patients within each cohort were invited, regardless of their access to mobile technology etc-however due to the design of the service this meant housebound patients could not attend. The project also did not systematically quantify or analyse medication changes made during the evaluation, limiting the ability to determine the relative contribution of education and treatment optimisation to improvements in HbA1c. In terms of patient and public involvement (PPI), there was no formal separate consultation stage of the programme design process, as the consultation process had consisted of getting informal feedback from service users who had been invited to attend similar groups pre-pandemic, which were more informal and conducted in a meeting room in the health centre itself. On reflection, it would have also been useful to obtain PPI feedback post-pandemic, as social attitudes to group education may have changed.

The association between implementation of the group education programme and subsequent improvements in glycaemic control suggests that the intervention contributed to observed outcomes. The group format provided structured disease education, peer discussion and supported initiation of injectable therapies, which are plausible mechanisms for behaviour change and treatment optimisation. This project suggests that group education has the potential to improve patient satisfaction and clinical outcomes when patients engage regularly. However, the findings suggest that a hybrid delivery model combining both group and individual approaches may be optimal to meet diverse patient needs and preferences.

A key learning point was that the patients initially identified as being most likely to benefit from the intervention were often among the least likely to attend. Many patients with multiple long-term conditions, mental health challenges, or other social complexities appeared to prefer individual consultations. The programme also highlighted the importance of peer support, which emerged as a particularly valued aspect of participation and may have contributed significantly to patient engagement and satisfaction.

Another key learning point from this project relates to data collection. Given the need to balance service delivery with existing clinical workloads, outcome measurement was intentionally focused on a limited number of parameters. Future iterations of the programme may benefit from the use of dedicated data capture tools, enabling a wider range of clinical, behavioural and patient-reported outcomes to be recorded and evaluated in a systematic manner, whilst maintaining the practicality of delivery within routine primary care.

## CONCLUSION

The project suggests that group education may improve confidence, knowledge and glycaemic outcomes for adults receiving injectable therapy for Type 2 diabetes.

Regular attendance appeared to be associated with greater improvements in HbA1c, whilst peer support emerged as the most valued feature of the intervention. However, attendance barriers remained substantial, particularly believed to be among individuals with multimorbidity and mental health conditions.

A hybrid care model combining group education with individual review may provide a more inclusive approach. Further evaluation at Primary Care Network level would help determine scalability and sustainability. As well as sharing the project results across PCN teams, learning has also been shared with the local Diabetes Specialist Nurse service. The model would be relatively easy to replicate where a suitable venue and dedicated clinical resources are available. Several smaller local groups may be more acceptable than a single larger programme covering a wider geographical area. Dissemination of these findings may support wider discussion regarding the role of community-based group education within contemporary diabetes care.

## Data Availability

All data produced in the present study are available upon reasonable request to the authors

## PATIENT AND PUBLIC INVOLVEMENT AND ENGAGEMENT

Patient attendees had previously been consulted for their feedback on a group education model during the previous group sessions prior to the COVID-19 pandemic, specifically stakeholders identified as people living with type 2 diabetes that were registered with the practice. There was an expression of preference for community-based education delivered locally, in preference to some of the education groups offered at the local hospital, or online. Feedback from participants was collected using satisfaction questionnaires and informal discussion during educational sessions. This feedback informed ongoing delivery of the sessions, including the allocation of time for group discussion and practical problem-solving. Findings from the project have been shared with participating patients, and their feedback will be used to guide future improvements to session content, accessibility and recruitment strategies.

## ACKNOWLEDGEMENTS

The authors would like to thank the participating patients, student nurses, administrative staff and community venue staff who supported the delivery of the programme. We would also like to thank Barney Jones for his advice and guidance regarding patient and public involvement and engagement (PPIE) during the development of this manuscript.

---

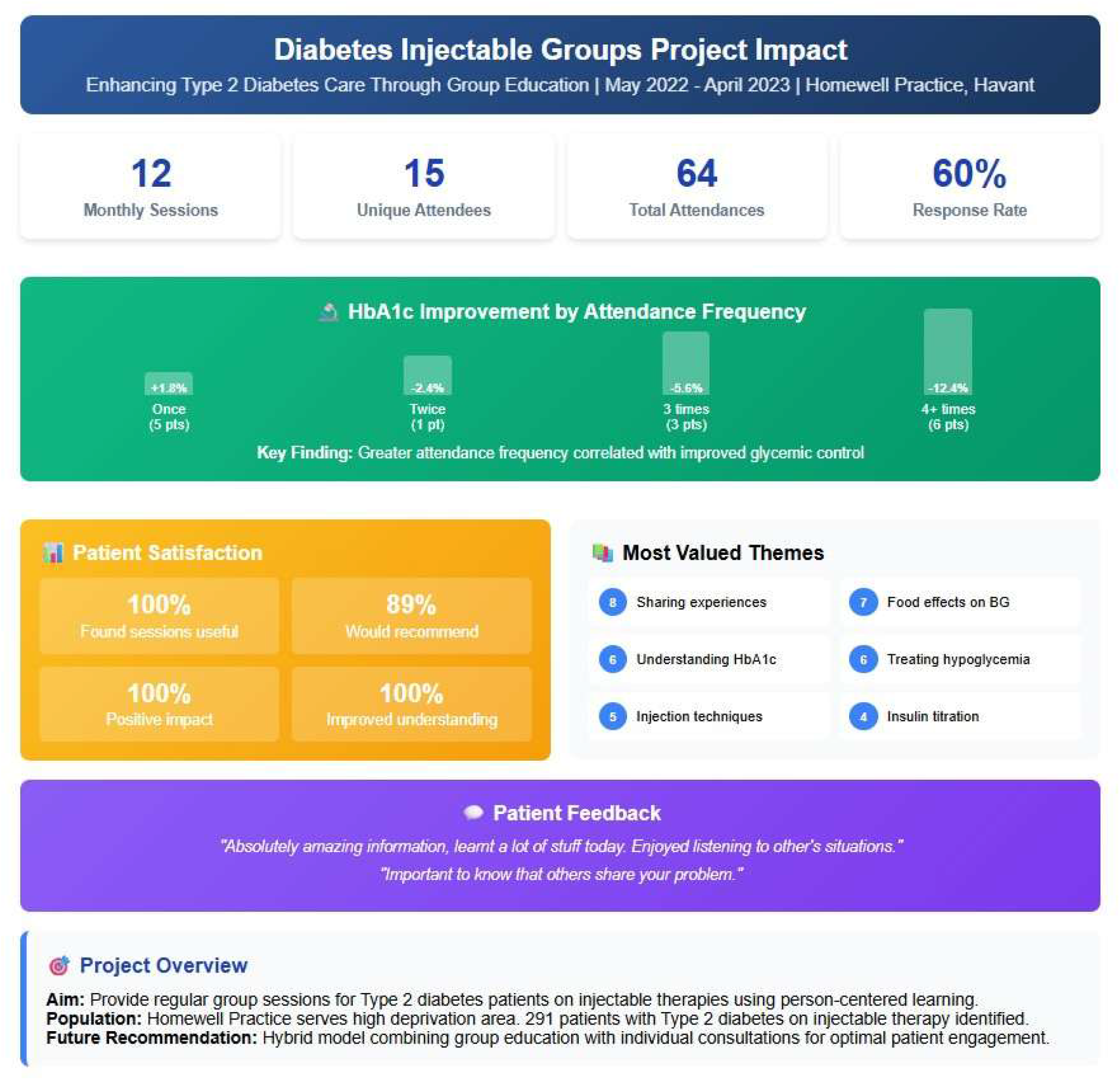

